# Distilling a High Precision Drug Adverse Effect Benchmark Using Wikipedia’s Wisdom of the Crowd

**DOI:** 10.1101/2021.11.01.21265741

**Authors:** Yonatan Bilu, Chen Yanover

**Author notes:** **Corresponding authors:** Yonatan Bilu, KI Research Institute, 11 Hazayit St, Kfar Malal, 4592000, Israel, Chen Yanover, KI Research Institute, 11 Hazayit St, Kfar Malal, 4592000, Israel.

## Abstract

**Background:** Large datasets of relational medical data, such as the adverse effects of drugs or vaccines, typically attain their large size, by relying on automatic, or semi-automatic, methods for generation. This often comes with a compromise on the precision of the generated data, which can be at least partially alleviated by having experts curate the data.

**Aim:** Since having experts review a large dataset can be costly and time consuming, here we suggest using Wikipedia for this task – that is, augment the automatic generation step by an automatic curation step based on the expert knowledge accumulated in Wikipedia.

**Methods:** To curate a dataset of adverse drug effects (ADEs), we suggest retrieving the Wikipedia page associated with the drug, and checking whether the ADE appears in the sections describing adverse effects. Drug indications, typically described in the opening paragraph of the page, are similarly filtered out.

**Results:** We use the method to curate two large adverse drug effect datasets and show that the obtained datasets have a much higher precision relative to their originating ones.

**Conclusions:** Algorithms which aim to infer drug-ADE relations, should at the very least be able to identify the “clear cut” cases. The high-precision benchmark constructed herein may therefore be a valuable resource for the evaluation of such algorithms.

## INTRODUCTION

The increased availability of real-world healthcare databases as well as the growing number of “big data” approaches to the analysis of such data hold the promise of alleviating some of the challenges inherent to assessing the safety and efficacy of medical intervention. Namely, alongside the prospective monitoring of long-term effects of interventions – such as drugs and vaccines – one can retrospectively analyze medical records in which such interventions are prevalent and try to identify conditions listed therein, which are caused by the intervention.

To support evaluation of causal methodologies in observational studies, comparative reference sets have been developed ^1,2^. Such sets, indicating the (direction of) effect of one drug ingredient compared to another on a given outcome, have been successfully used to assess the performance of multiple comparative effectiveness methods, e.g. Ryan et al.^3^. However, evaluation of some tasks – importantly, estimating the efficacy of a vaccine or the safety of drugs –requires reference sets that compare the effect of a treatment on an outcome, relative to no-treatment (or placebo).

Previous efforts to generate drug vs placebo reference sets relied on manual curation of drug-adverse event pairs, resulting in limited, few hundred pair sets^1^. Alternatively, one can obtain much larger sets from knowledgebases, such as the SIDER dataset^4^, which extracted adverse drug effects (ADEs) from public documents and package inserts using natural language processing (NLP) methods, or OFFSIDES^5^ dataset, which is based on the FDA Adverse Event Reporting System. However, the obtained lists typically mix rare and common ADEs alongside those caused by misuse or abuse of drugs^6^.

Indeed, determining what constitutes a “true” effect of a drug may be unclear, and from a functional perspective – may depend on the context in which this question is asked, as well as the drug use patterns and prevalence of the adverse effect. For example, in comparing SIDER and OFFSIDES, Cheng et al.^7^ found relatively little overlap between them (7,741 common pairs, out of 418,532 OFFSIDES pairs and 120,236 SIDER pairs) suggesting that it is common for ADEs described in package inserts not to be reported to FAERS, and vice versa. Rare yet severe side effects may be of great interest to patients and clinicians, even if they were not detected during clinical trials, and, conversely, drug manufacturers may opt to be cautious and include such effects in the package insert, even if they are rarely reported.

However, in the context of prediction (or estimation) algorithms – in particular, those which infer causal relations from electronic health records (EHRs) - it is important to differentiate between effects which can be clearly attributed to a properly used drug, and those where the attribution is unclear. While an ideal machine-learning algorithm would detect all “true” side effects, its success is limited by the data it is trained on, and our confidence in its evaluation is dependent on our confidence in the veracity of the data used for evaluation. Hence, in this context, it is useful to construct a benchmark of ADEs which are clearly caused by a drug. That is, not an exhaustive list of all “true” ADEs, but rather a benchmark of ADEs that we expect any good prediction algorithm to detect with high accuracy. The approach we explore here to create such a benchmark is to start from an expansive list of ADEs, and distil from it those ADEs which are frequently encountered in practice.

One way to do this is for expert medical professionals to manually curate a large dataset and pinpoint, based on their medical experience, adverse effects commonly observed in practice. However, this can be potentially biased, and is usually costly and time consuming – especially if one wants to integrate the experiences of several medical professionals. An alternative, which encapsulates the same motivation, is to cross-reference the adverse effects listed in a dataset, with those mentioned in Wikipedia. Indeed, Wikipedia articles often reflect the aggregated knowledge of multiple editors, many of which are well versed in the field on which they write.

Here we apply NLP methods on Wikipedia drug articles to verify SIDER and OFFSIDES ADEs and come up with more concise lists (*Figure 1*). We demonstrate the advantage that the resultant set provides for evaluating prediction algorithms, by showing that a larger fraction of the ADEs therein overlap those obtained from published clinical trials as well as manually-constructed ones. We hope that our distilled lists (provided in the supplementary) serve as a benchmark to systematically test placebo-based study designs and methodologies.

**Figure 1:**
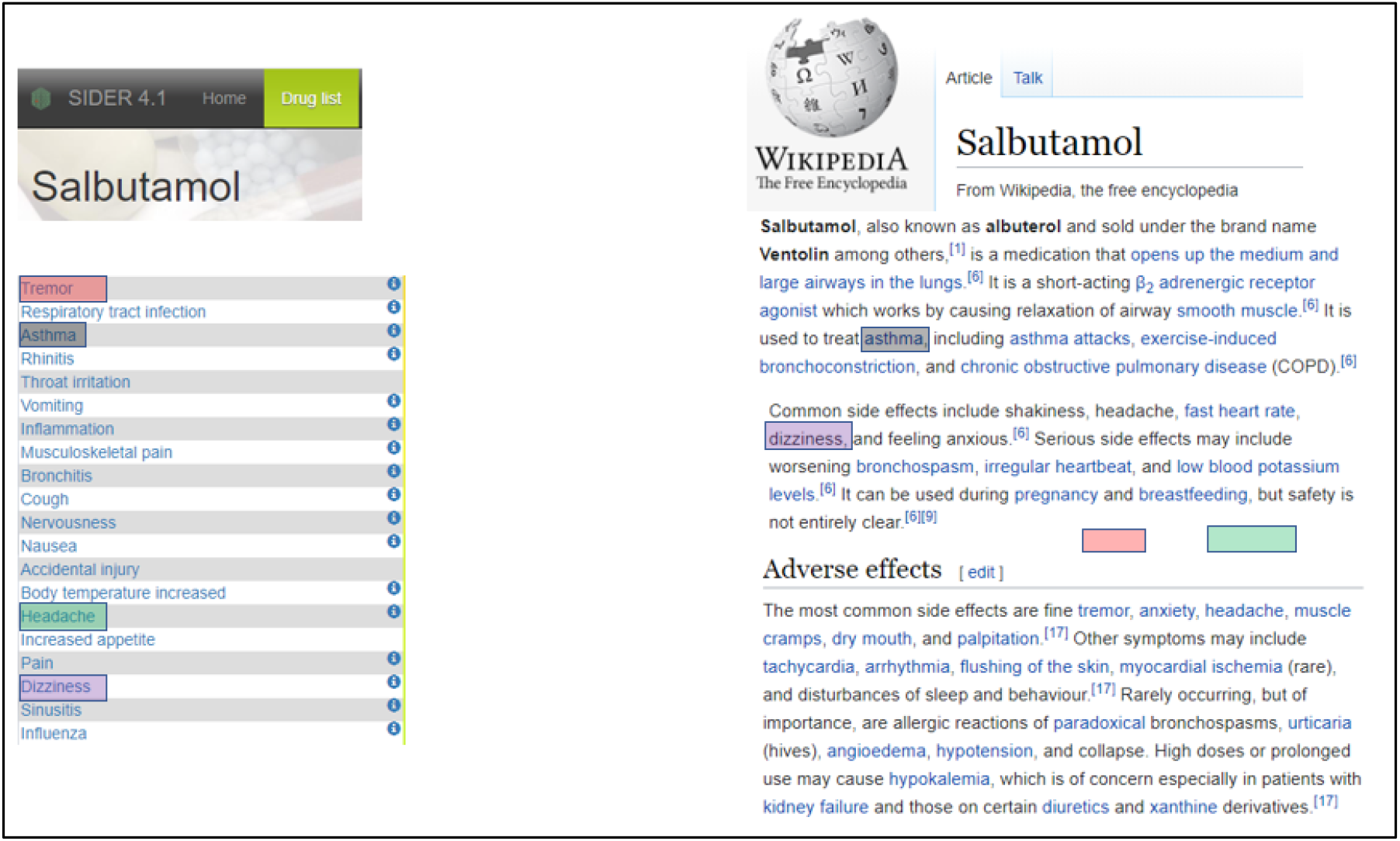
Snippet of the of the ADEs listed for the drug Salbutamol in SIDER (left), and the relevant sections from the Wikipedia page for this drug (right). Matching ADEs are highlighted. Since “asthma” appears in the first paragraph of the article, it is considered an indication, and not verified. Note that some of the ADEs listed in SIDER are unlikely to be supported by EHRs (e.g., influenza), exemplifying the need for “distilling” this list.

## METHODS

### Data

We consider two large datasets of drugs adverse effects, SIDER (version 4.1)^4^ and OFFSIDES (version 0.1)^5^, the former consisting of 309,849 drug-ADE pairs (with repetitions), and the latter of 3,206,558 pairs. In addition, in OFFSIDES each pair is annotated with a proportional reporting ratio (PRR), and SIDER provides a list of drug-indication pairs. We use this data to filter the datasets, as described in the supplementary methods section), and end up with 129,513 pairs in the SIDER datasets (1344 drugs, 3753 ADEs), and 1,051,942 in the OFFSIDES one (2381 drugs, 9957 side effects). There are 41,332 pairs common to both sets (985 drugs, 2497 side effects).

In total, these datasets refer to 2724 different drugs. We attempted to retrieve their associated Wikipedia articles, using the Wikipedia API, on August 9^th^, 2021. For 233 (8.5%) of them, no article was found.

### Distilling by Wikipedia

To identify ADEs mentioned in a drug’s Wikipedia article we rely on two simplifying observations in our setting: (1) We do not need to identify all ADEs, only to determine which of those mentioned in SIDER or OFFSIDES also appear in the Wikipedia article; and (2) such articles usually mention adverse effects either in the initial summary, or in a dedicated section.

Given a name of a drug, we retrieve the corresponding Wikipedia article, extract the relevant sections and match ADEs from SIDER and OFFSIDES as described in the supplementary methods. ADEs which are mentioned in the first paragraph of the article are considered indications, and are discarded. In addition, where possible, the frequency of an ADE is deduced by keywords appearing in the sentence in which it was mentioned. Drugs for which no Wikipedia article is found, or for which the Wikipedia article does not contain a relevant section, are discarded from the analysis. In total, 13,570 pairs were “verified” this way.

### Distilling by PRR

We interpret the PRR of a drug-ADE pair as a confidence score in the drug indeed causing the ADE, and accordingly employ it in two ways. First, we examine it as an alternative to distilling by Wikipedia. Specifically, if for given a drug, the Wikipedia distillation verifies *k* of the ADEs listed in OFFSIDES, then we compare these *k* ADEs to the *k* ADEs with the highest PRR lower bound computed for this drug. We denote this list OFFSIDES-top.

Second, we consider drug-ADE pairs with higher PRR as more likely to represent true ADEs. We limit our analysis to pairs where the drug has a Wikipedia article with a relevant section (779,064 pairs in total). For a given integer *n*, we define OFFSIDES_n_ as the *n* pairs with the highest PRR lower bound score, and OFFSIDES-Distilled_n_ as the fraction of these pairs which are verified by Wikipedia. Assuming that for smaller values of *n* the fraction of true ADEs is higher, and that the distillation process tends to identify these pairs, we expected that OFFSIDES-Distilled_n_ is high for small values of *n*, declining as *n* grows, as suggested in the Results.

### Evaluation

We first evaluated the sensitivity and specificity of the distilled adverse event set, compared to the original one, on a ground truth set derived from clinical trials, using the code of Steinberg et al.^2^ and requiring that one of the treatment arms is placebo (leaving all other parameters unchanged). The clinical trial set included 412 drug-ADE pairs. Additionally, we examined how well aligned these lists are with the reference list of Voss et al.^8^, which lists 208 drug-ADE pairs for a nine ADEs. For all evaluations, we report precision, recall, and F1 score – the harmonic mean of these two measures.

We assume that in both sets, the drug-ADE pairs are of the type we are interested – the ADE is clearly caused by the drug, and should therefore be classified as such by a good prediction algorithm. Hence, our goal is to show that distilling by Wikipedia yields a concise list of drug-ADE-pairs where a relative a high proportion overlaps these sets.

We note that simply distilling a list at random is likely to reduce the F1 score. Suppose that a dataset achieves a precision of *p* and a recall of *r* when compared to some benchmark. Suppose that we pick at random an *f* fraction of the pairs in the dataset as our distilled set. Then the precision for this subset is expected to be *p* (as before), but the recall is expected to be *rf*. Since the F1 score is the (harmonic) mean of these two measures, and *rf < r*, the F1 of the randomly distilled set is expected to be lower than that of the original set.

## RESULTS

### Distilling statistics

Among the 129,513 pairs from SIDER, 9,386 were Wikipedia-verified (781 drugs, 826 ADEs), and from the 1,051,942 OFFSIDES pairs 9177 were Wikipedia-verified (1002 drugs, 1152 ADEs). In other words, distilling SIDER leaves 7.2% of the initial set, and distilling OFFSIDES leaves less than 1%. However, the overlap between the two distilled sets is 4,993 pairs, that is, 12.1% of the 41,332 pairs common to both SIDER and OFFSIDES. That is, considering the overlap between SIDER and OFFSIDES as more likely to contain reliable pairs, we see that indeed distilling identifies a larger fraction of pairs therein.

The distilled set maintains a high level of diversity in the sense that there is no single drug, or small subset of drugs, which dominates the distilled set – the most common drug there, zuclopenthixol, appears in less than 0.7% of the pairs. By contrast, some side effects, such as nausea and headache, are relatively common, appearing in 3.5% and 2.8% of the pairs respectively.

### Comparison to gold standards

The much smaller distilled lists obtain, expectedly, lower recall but its precision and F1 score are higher, in the comparisons with both ground truth sets (**Error! Reference source not found**.). Results are presented both for all pairs which were verified by Wikipedia, and – for the Clinical Trials benchmark - a subset of them, for which the frequency of the ADE was indicated, in the drug’s Wikipedia page, to be common (for the benchmark of Voss et al, initially there is very little overlap with either SIDER or OFFSIDES, and after filtering only those indicated to be common, the precision and recall are 0).

**Table 1.** Evaluation of SIDER, OFFSIDES and their distilled set, vs. two ground truths, derived from Clinical Trials^*2*^ and manual curation^*8*^. All measures are micro-averaged over drug-ADE pairs.

| Ground truth | Reference Set | Precision | Recall | F1 |
| --- | --- | --- | --- | --- |
| Clinical Trials | SIDER | 0.14 | 0.89 | 0.23 |
|  | SIDER-Distilled | 0.86 | 0.70 | 0.75 |
|  | SIDER-Distilled common | 0.96 | 0.31 | 0.42 |
|  | OFFSIDES | 0.04 | 0.62 | 0.08 |
|  | OFFSIDES-top | 0.08 | 0.07 | 0.07 |
|  | OFFSIDES-Distilled | 0.70 | 0.50 | 0.55 |
|  | OFFSIDES-Distilled common | 0.87 | 0.21 | 0.30 |
| Voss et al. | SIDER | 0.003 | 0.22 | 0.005 |
|  | SIDER-Distilled | 0.014 | 0.10 | 0.021 |
|  | OFFSIDES | 0.0006 | 0.13 | 0.001 |
|  | OFFSIDES-top | 0.002 | 0.03 | 0.004 |
|  | OFFSIDES-Distilled | 0.008 | 0.08 | 0.014 |

### Comparison to high-PRR OFFSIDES pairs

Looking at subsets of OFFSIDES drug-ADE pairs defined by their PRR score, we see that for a high PRR threshold a relatively large portion of the pairs is considered verified by the process suggested here (Figure 2). For example, if we consider the 100 pairs with highest PRR score, some 10% of them are verified. This number decreases some ten folds as the threshold on PRR is lowered. However, overall, there seems to be no correlation between the PRR score and a drug-ADE pair being verified (Pearson Correlation; r=0.004).

**Figure 2.**
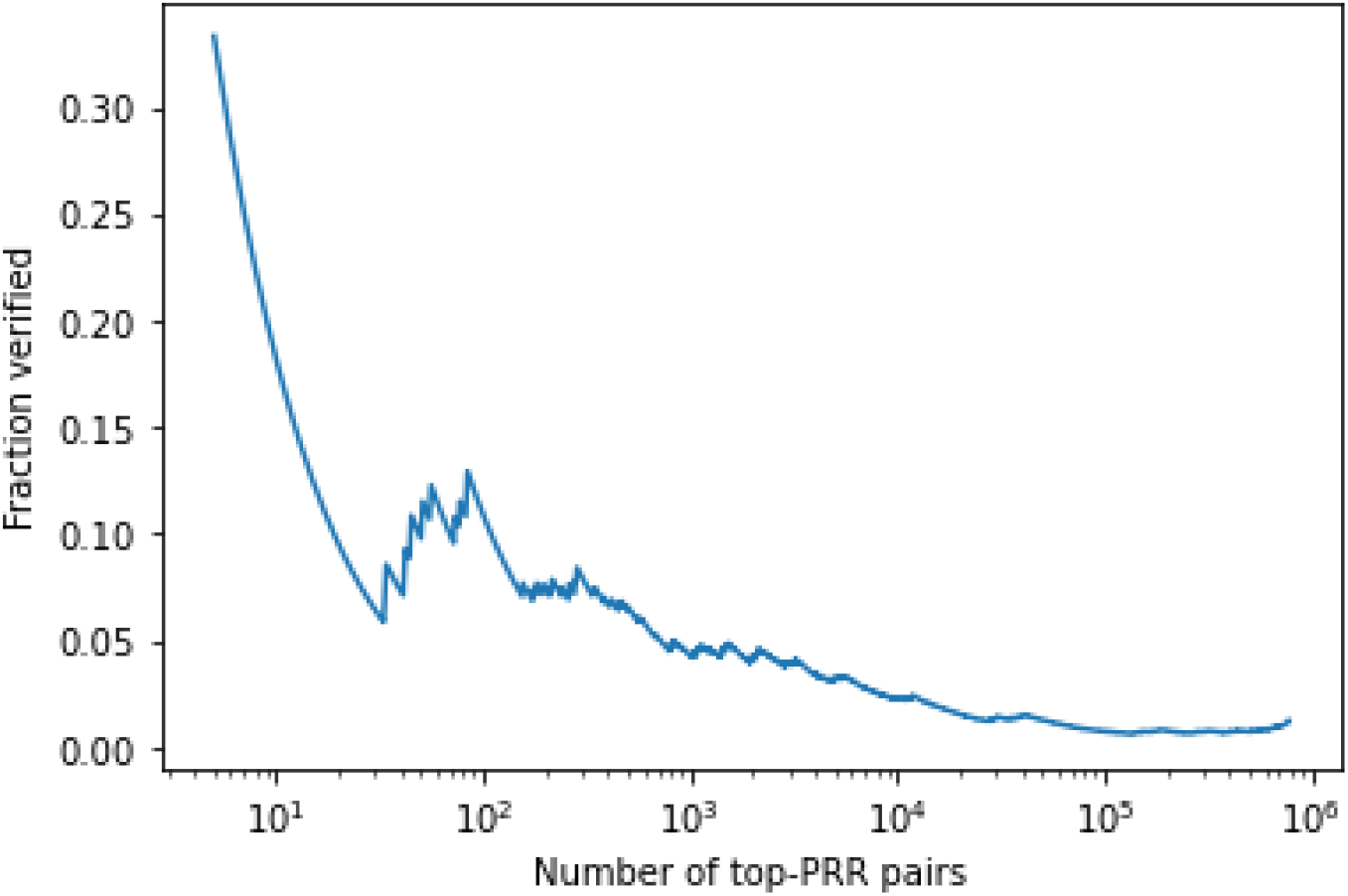
Fraction of verified OFFSIDES drug-ADE pairs by number of PRR-sorted pairs. Overall, including more top pairs, that is, using a more relaxed PRR threshold, decreases the fraction of verified pairs, suggesting that our distilling process prefers pairs with higher confidence.

## DISCUSSION

We present an automatically-curated dataset of drug-ADEs, focusing on positive examples. This dataset can be useful not only as a benchmark for the prediction of ADEs from EHRs, but also for general prediction and inference methods. Exhibiting good performance on this dataset is indicative of the method’s power, and suggests that it could work well when performing other tasks, for which curated data is scarce or unavailable – such as the adverse effects of vaccines.

Importantly, this dataset is oriented towards precision – it does not aim to capture all ADEs that a drug may cause. Rather, it lists a subset of these ADEs, for which confidence is high. Hence, evaluating a model versus this benchmark should be done accordingly.

Relying on massive knowledgebases is but one approach to the question of how to calibrate and validate causal inference methods. For example, the common practice in the OHDSI community, which promotes observational studies and develops tools to conduct them, is to build a reference set in two steps. First, to identify negative controls (examples of drug-ADE pairs which should not be causally related), either manually or from literature, product labels, and spontaneous reports^8,9^; then, derive synthetic positive controls from the obtained negative controls. We note, however, that the resulting list of negative controls can be distilled using an approach similar to the one outlined here.

In future work, we aim to evaluate this dataset versus predictions deduced from EHRs. Initial research in this direction, using OHDSI’s SelfControlledCaseSeries package^10^, and naively labeling drug-ADE pairs according to the sign of their log ratio scores, revealed two inherent problems: (1) In high-scoring pairs, the effect is almost always an indication rather than an ADE; and (2) effects common in the data tend to get a high score, even when we could find no external corroboration for them being an ADE. Addressing these problems will also allow to evaluate a set of negative controls.

The NLP techniques used herein are rather naïve. Specifically, when matching an ADE to Wikipedia, the current approach may miss ADEs which are named differently in Wikipedia and in SIDER or OFFSIDES. Initial exploration into more sophisticated matchings, such as stemming or lemmatizing terms before matching, or transforming terms into an embedded representation and considering distances in the embedding space^11,12^, have so far not but fruitful: Considering stemmed and lemmatized versions of the terms did not extend coverage significantly, and relying on embedded representations introduced many erroneous matches.

Finally, we believe that the method developed here may have broader use. We showed that a rather naïve use of Wikipedia to curate medical data can greatly improve the accuracy of the results. It is likely that this can be extended to other types of data.

## Supporting information

Distilled drug-ADE list

## Data Availability

All data produced in the present study are available as supplementary materials.

## AUTHORS’ CONTRIBUTIONS

YB and CY conceived the study; acquired, processed, analyzed, and interpreted the data; and wrote the manuscript.

## SUMMARY TABLE

### Already known on this topic

- Evaluation of algorithms that estimate the safety of medical interventions (e.g., drugs or vaccines) from observational data requires a benchmark set of adverse effects that can be identified in electronic health records
- Large knowledgebases of adverse drug effects (ADEs), extracted from various sources of information, include adverse effects of varying prevalence and severity, caused by proper use or misuse of medications; and these, often, cannot be identified in routinely collected medical data

Manual curation of large datasets by domain experts is costly and time consuming, and typically results in small benchmark sets.

### What this study adds

- We apply natural language processing algorithms on Wikipedia, as a crowd-sourced panel of professionals, to automatically distill ADE lists from large datasets and demonstrate the higher precision of these sets compared to the original ones
- The obtained distilled lists may be a valuable resource for the evaluation of prediction and estimation algorithms

## APPENDIX: SUPPLEMENTARY METHODS

### Processing of SIDER and OFFSIDES

SIDER lists a total of 309,849 drug-ADE pairs (with repetitions), covering a total of 1430 drugs and 6123 ADEs. OFFSIDES lists a total of 3,206,558 pairs, 2730 drugs and 14,544 ADEs. In addition, in OFFSIDES each pair is annotated with a proportional reporting ratio (PRR; the ratio between the frequency reported for the ADE in those taking the drug, and in those taking other drugs).

For both datasets, we convert drug codes to ATC level5 codes. For ADEs, which in both datasets employ the MedDRA vocabulary, we take only those from hierarchy level PT (preferred term) or LLT (lower-level term). In the latter case, we convert the ADE to the parent PT. In the case of OFFSIDES we limit the analysis to pairs where the PRR is listed, and bigger than 1 + the listed PRR error term.

The SIDER dataset provides, in addition to ADEs, a list of drug-indication pairs. We remove from both datasets drug-ADE pairs when the ADE is also listed as an indication for the drug. After all this filtering, and the removal of duplicates, we are left with 129,513 pairs in the SIDER datasets (1344 drugs, 3753 ADEs), and 1,051,942 in the OFFSIDES one (2381 drugs, 9957 side effects). There are 41,332 pairs common to both sets (985 drugs, 2497 side effects).

### Distilling by Wikipedia

Given a name of a drug, we retrieve the corresponding Wikipedia article, and process it as follows: we search for sections whose title contains the phrases: “side effect” or “adverse”. If such a section is found, it is parsed into subsections and then into sentences. Subsections are discarded if their title contains any of the words - ‘tolerance’, ‘dependence’, ‘withdrawal’, ‘interaction’, ‘overdose’, ‘discontinuation’, since we are interested in adverse effects with respect to placebo. In addition, the opening summary section is extracted, and parsed into paragraphs and sentences. Drugs for which no Wikipedia article is found, or for which the Wikipedia article does not contain a relevant section, are discarded from the analysis.

An ADE mentioned in SIDER or OFFSIDES is considered “verified” by Wikipedia if it appears in one of the sentences extracted as above. We examine only ADEs which belong to the “preferred term” hierarchy level in MedDRA, and consider an ADE as “verified” also if one of its “lower-level terms” in the hierarchy is mentioned. For ADEs which are multi-word expressions we allow any word order, and ignore stop-words in both the name of the ADE and the Wikipedia text. If a condition appears in the first section of the article, we suspect that it’s an indication and not an ADE, and so do not verify it. In total, 13,570 pairs were “verified” this way, of which 8609 were matched to the exact term, and 4961 to “lower-level term”.

In addition, where possible, the frequency of an ADE is deduced by keywords appearing in the sentence in which it was mentioned, or in the subsection’s title. Namely, we consider an ADE as common, if one of the following keywords is mentioned – common, commonly, frequent, frequently, often; we consider it uncommon based on the keywords – infrequent, uncommon, and less common; and we consider it rare by the keywords rare or rarely.

We discard ADEs which are mentioned in the first paragraph of the Wikipedia article’s summary section – since this paragraph typically mentions indication. The natural language processing done here made use of python’s nltk^13^ package.

