## Supplementary material for "Distilling a High Precision Drug Adverse Effect Benchmark Using Wikipedia’s Wisdom of the Crowd": Distilled drug-ADE list: README.txt

The file "distilled drug-ADEs.csv" contains the data generated and analyzed in "Wikipedia Drug Safety Advisory Committee: Distilling a Drug Adverse Effect Reference Set Using Wisdom of the Crowd"

The columns of the file are as follows:

drug - the ATC-5 name of the drug ingredient
ADE - the MedDRA name of the adverse drug effect
sider - whether or not the drug-ADE pair appears in SIDER
offsides - whether or not the drug-ADE pair appears in OFFSIDES
prr_lb - for pairs appearing in OFFSIDES, a lower bound on the PRR; the listed score minus the listed error term.
wiki_status - whether or not the ADE was found in the relevant sections of the Wikipedia page of the drug. Blank if no relevant article was found.
wiki_indication - whether or not the ADE was found in the first paragraph of the Wikipedia page of the drug.
wiki_freq - a frequency indicating keyword that was found in same sentence as the ADE, if one was found. If the ADE appears in more than one sentence, more than one keyword may appear.
wiki_term - the actual term found in Wikipedia and matched to the ADE. This differs from the ADE if the matching was to a MedDRA LLT.
 
